# A neuroimaging measure to capture heterogeneous patterns of atrophy in Parkinson’s disease and dementia with Lewy bodies

**DOI:** 10.1101/2023.08.01.23293480

**Authors:** R Bhome, S Verdi, SA Martin, N Hannaway, I Dobreva, NP Oxtoby, Castro Leal G, S Rutherford, AF Marquand, RS Weil, JH Cole

## Abstract

**INTRODUCTION:** Parkinson’s disease (PD) and Dementia with Lewy bodies (DLB) show heterogeneous brain atrophy patterns and common group-average analyses are limited in capturing individual differences. Neuroanatomical normative modelling overcomes this by comparing individuals to a large reference cohort.

**METHODS:** We generated z-scores from T1w-MRI scans for each participant (108 PD; 61 DLB) relative to normative regional cortical thickness and subcortical volumes, modelled in a reference cohort (n=58,836). Outliers (z<-1.96) were aggregated across 169 brain regions per participant. We examined total outlier counts between high versus low visual performance in PD; and PD versus DLB; and tested associations between these and cognition.

**RESULTS:** We found greater total outlier counts in PD poor visual performers, compared to high; and in DLB versus PD. Outlier counts were associated with global cognition in DLB, and visuoperception in PD.

**DISCUSSION:** Neuroanatomical normative modelling shows promise as a clinically informative technique in PD and DLB.

## Introduction

Cognitive impairment is a core diagnostic feature of Dementia with Lewy bodies (DLB) [1] and is common in Parkinson’s disease (PD) where almost half of patients develop dementia within ten years’ of diagnosis [2]. Conventional neuroimaging studies measuring cortical thickness and grey matter volume in DLB and PD patients, analysed at group level, have yielded heterogeneous findings [3, 4]. Previous work has failed to identify a consistent pattern of atrophy that predicts future cognitive decline [5–7] and correlates with symptom severity [4], thereby limiting the value of conventional neuroimaging measures as biomarkers.

A key challenge of studying neurodegenerative diseases at group level is between-subject heterogeneity, which results from intrinsic biological differences as well as psychosocial and environmental factors independent of the presence of disease [8]. This has significant implications for conventional case-control neuroimaging studies that compare group means, which only allow inferences to be made for the ‘average subject’ and treat between-subject variability as noise [9].

To improve our knowledge of the neural basis of neurodegenerative disorders such as PD and DLB, there is a need to understand between-patient heterogeneity. Neuroanatomical normative modelling is a recently established neuroimaging analysis framework that precisely maps individual patterns of variation from the expected norm for a given neuroimaging measure [9–11]. A recent endeavour has developed a robust neuroanatomical normative reference model from which individual-level predictions can be made [12]. Lifespan trajectories of cortical thickness and subcortical volumes were modelled using Bayesian Linear Regression based on a reference cohort of 58,836 healthy participants, adjusting for age, sex and site. Leveraging this information, a new individual’s cortical and subcortical data can be plotted within each normative distribution, to quantify the extent they deviate from expected patterns. Predetermined values can be used to binarise z-scores, to quantify how many, and in which regions the individual is an outlier. The number of outliers can be aggregated to provide the total outlier count, a measure of overall neurodegeneration at individual-level. This may be a more effective biomarker of brain health, particularly in Lewy body disorders where patterns of atrophy are highly variable [3, 4].

Neuroanatomical normative modelling has recently been successfully applied in Alzheimer’s disease (AD) [13–15]. As expected, patients with AD had more outlier regions than mild cognitive impairment (MCI) and healthy control groups [14], though neuroanatomical patterns of outliers were variable even within AD groups [14, 15]. Importantly, total outlier count correlated with poorer cognitive performance, fluid biomarker-measures of Alzheimer’s pathology, and predicted future conversion from MCI to dementia [13, 14]. Given that neuroimaging measures in AD may be more neuroanatomically homogenous than Lewy body disorders [16, 17], this approach may have even greater utility in Parkinson’s and DLB.

Here, we leveraged this technique to investigate heterogeneity in Lewy body diseases and evaluate the potential of neuroanatomical normative modelling to provide useful measures of disease severity. In PD, previous work has shown that visual performance predicts future cognitive decline, with the distinction between high and poor visual performance found to represent risk of future dementia in PD [18, 19]. Here, we a) investigated differences in total outlier count between high and poor visual performers with PD, and compared it to conventional cortical thickness analysis; b) identified individual patterns of outliers in cortical thickness and subcortical volumes in PD and DLB; c) compared patterns of dissimilarity between PD participants with high versus poor visual function; and between PD and DLB participants, and d) evaluated whether total outlier count correlated with cognitive severity in PD and DLB. We hypothesised that there would be significant differences in total number of regional outliers between high and poor visual performance PD groups; but that conventional case-control analysis of cortical thicknesses would not identify group differences. We further hypothesised greater dissimilarity in individual patients for low versus high visual performers in PD; and for DLB compared to PD. Finally, we predicted that greater total outlier count would be significantly associated with poorer cognitive performance in PD and DLB.

## Methods

### Participants

Structural T1w-MRI data from two sites were used. The first site, at the Wellcome Centre for Human Neuroimaging, UCL, included 108 participants with PD, 36 with DLB and 38 healthy controls, who are part of the Vision in Parkinson’s disease study (PI: Dr Rimona Weil, NRES Queen Square Ethics Committee reference 15/LO/00476). The second site was the pseudoanonymised Alzheimer’s Disease Research Center (ADRC) “8361” which contributes data to the National Alzheimer’s Coordinating Center (NACC) database [20], and included 25 participants with DLB and 127 healthy controls. Participants from the UCL site were recruited from the National Hospital for Neurology and Neurosurgery outpatient clinics and affiliated hospitals, or from patient and carer group bodies such as the Lewy Body Society and Rare Dementia Support. They were diagnosed as having PD or DLB if they satisfied the Queen Square Brain Bank PD diagnostic criteria [21] or the Dementia with Lewy Bodies Consortium Criteria [1] respectively, with exclusions if they had a history of traumatic brain injury, major co-morbid psychiatric or confounding neurological disorder. Additionally, all UCL participants were assessed by a neurologist (RSW) to make a positive diagnosis of PD or DLB and to identify clinical features or signs on neurological examination that could be suggestive of an alternative atypical parkinsonian syndrome such as progressive supranuclear palsy or multisystem atrophy (e.g., supranuclear gaze palsy, cerebellar signs, prominent dysphagia, early and prominent urinary symptoms, severe autonomic involvement). Controls were recruited from spouses of patients taking part in the study or the UCL Institute of Cognitive Neuroscience volunteer databases.

Participants from site “8361” were included and classed as having DLB if they had a structural MRI scan and met the following criteria based on clinical descriptors available in the NACC data file (June 2022 data freeze): 1) dementia diagnosis; 2) Primary, contributing, or non-contributing cause of cognitive impairment - Lewy body disease; 3) not classed as MCI; and 4) the absence of a diagnosis of Parkinson’s disease.

### Clinical assessment

96 of the PD participants at the UCL site had been further divided into high (n=64) and low (n=32) visual performers based on their performance on two computerised visual tasks, the biological motion task and the Cats-and-dogs task, which have been described previously [18, 22, 23] and have been shown to predict dementia and poor outcomes in Parkinson’s [18, 19, 24]. In this way the low visual performers are thought to represent an at-risk group for conversion to Parkinson’s dementia.

Clinical assessment included detailed neuropsychometric testing and disease specific measures of clinical severity. For this study, we primarily focused on a global measure of cognition: the Montreal Cognitive Assessment (MoCA) score [25], which is widely used clinically as a sensitive measure of global cognitive function in Parkinson’s disease [26, 27] as well as a composite cognitive score [18, 19]. This composite score combined measures across five cognitive domains, using the averaged z-score for each of the following cognitive tests: Stroop colour time [28]; verbal fluency [29]; word recognition [30]; Hooper Visual Organisation Test [31] and MoCA [25]. Additionally, as visuo-perceptual ability is usually affected early in DLB [1], we also specifically examined performance in a visuo-perceptual cognitive test, the Hooper Visual Organisation Test [31].

Disease specific measures included the Movement Disorder Society Unified Parkinson’s Disease Rating Scale (MDS-UPDRS) that measures impairment and disease severity in both motor and non-motor domains [32], part III of the MDS-UPDRS (MDS-UPDRS-III) to assess motor function [32], the University of Miami Hallucinations Questionnaire (UM-PDHQ) to evaluate hallucinations [33] and depression severity was measured with the Hospital Anxiety and Depression Scale (HADS) [34].

### MRI acquisition and processing

Structural T1w-MRI scans at UCL were acquired on a single 3T Siemens Magnetom Prisma scanner with a 64-channel head coil. Structural magnetisation prepared rapid acquisition gradient echo (MPRAGE) data were acquired using the following parameters: 1×1×1mm voxel, TE=3.34ms, TR=2530ms, flip angle=7°. Acquisition time for MPRAGE was approximately 9 minutes. Structural T1w-MRI scans from NACC ADRC “ 8361” were acquired on 1.5T scanners manufactured by GE. Further information on individual scan parameters are available via the NACC database.

The “recon-all” function in FreeSurfer v6.0.0 (http://www.freesurfer.net) was used to process all MRI data from both the UCL and NACC sites. Cortical thickness values of the Destrieux parcellation (lh.aparc.a2009s.stats, rh.aparc.a2009s.stats) [35] and subcortical volumes (aseg.stats) were extracted. Cortical parcellations and subcortical segmentations were manually quality controlled by visually inspecting these segmentations superimposed on the corresponding structural T1-weighted image by a researcher who was blind to clinical status.

### Reference normative dataset

Rutherford and colleagues [12] modelled normative lifespan trajectories for cortical thicknesses across 148 regions using the Destrieux parcellation and subcortical volumes derived from Freesurfer (v6.0.0), using a warped Bayesian Linear Regression with age and sex as covariates, and also accounting for site differences [36]. The employment of Bayesian linear regression with likelihood warping allows accurate modelling of non-Gaussian effects and upscaling of normative models to large cohorts [37]. Their reference cohort comprised 58,836 subjects from 82 sites and this large number of participants across multiple sites further strengthened the reliability of the distribution estimates.

### Applying neuroimaging normative modelling to the study data

The reference normative model was recalibrated to the specific datasets used in our study by using an adapted transfer learning approach [38]. This involved inputting healthy controls’ data from the two sites in our study into the reference normative model to generate stable parameters for cortical thicknesses and subcortical volumes, which the individuals within the disease groups could then be validly measured against. To do this, z-scores were generated for each individual with either DLB or PD for each cortical region and subcortical structure, relative to the recalibrated reference values. All modelling steps were performed using PCNToolkit (v0.20) and by applying freely available code via Google Colab [12].

### Statistical Analysis

#### Total outlier count

From the z-scores for each cortical region and 21 subcortical structures (accumbens, amygdala, caudate, cerebellar cortex, cerebellar white matter, hippocampus, pallidum, putamen, thalamus, ventral diencephalon, all bilaterally, and the brain stem) generated from the normative modelling pipeline described above, outliers were defined as z-scores < −1.96. The total number of outliers across the 169 regions and structures was summed for each participant to provide a single metric per participant known as the total outlier count. Linear regressions, correcting for age and sex, were used to test for group differences in total outlier count between high and low visual performers with PD as well as between DLB and PD. Further, subgroup analyses compared DLB participants at the UCL site with the NACC site and PD and DLB participants only at the UCL site. Additionally, group comparisons for proportion of outliers at each region were conducted using a Mann-Whitney U test and corrected for multiple comparisons using the False Discovery Rate (FDR).

### Measuring dissimilarity within and between groups

The Hamming distance metric is widely used in information theory and reflects the dissimilarity between two strings of equal length. At each point on the strings, a distance of 1 is assigned if the symbols are different and a distance of 0 if the symbols are the same. This is then summed across the length of the string to give the total Hamming distance [39]. This metric was applied to the vector of binarised z-scores for outliers across 169 brain regions.

Participants were compared pairwise within groups, so had *n-1* Hamming distance scores ranging from 0-169, where *n* is the number of participants in their group. The median Hamming distance score for each participant was then calculated and between-group comparisons were made using the median Hamming distances, using a Mann-Whitney U test.

To qualitatively visualise spatial patterns of cortical thickness outliers per group, the proportion of participants within each group who were outliers based on their z-score (<-1.96) for each cortical region was calculated. This was mapped using the Destrieux atlas via the R package ggseg [40].

### Association of total outlier count and clinical features

Linear regressions adjusting for age and sex were used to test the association between total outlier count and a composite cognitive score (see above), MoCA and visuoperception, measured using the Hooper Visual Organisation Test. In a secondary analysis, we also tested the association with disease specific measures including a global measure of severity (MDS-UPDRS), motor symptoms (MDS-UPDRS-III), hallucination severity (UM-PDHQ) and depression score (HADS). The association was tested in the PD and DLB groups separately, and for the DLB group we only included data from UCL where clinical severity data had been robustly collected.

Statistical analyses for normative modelling data were performed in R (v4.2.2).

### Potential outliers in total outlier count measure

There was one participant from the PD group and one from each of the DLB groups at the UCL and NACC site, who had a much higher total outlier count than the other participants in the group (45, 50 and 53, respectively). Their brain imaging was initially quality controlled by one of the authors, RB, and then further scrutinised by additional authors (RSW and JHC). The participants were deemed to not warrant exclusion as there was not a clear acquisition or processing error leading to the total outlier count. Further, the individuals’ total outlier counts are biologically plausible and where the authors (RB and RSW) had clinically examined the two participants at the UCL site, their diagnoses were unambiguous based on clinical examination and neuroimaging. We present results with and without the inclusion of these participants in analyses below.

### Conventional cortical thickness analysis

We also evaluated whether there were regional differences in cortical thickness at a group level between high and low visual performers with PD and between PD and DLB using a conventional General Linear Model (GLM) (Freesurfer Version 6.0). This incorporated Monte Carlo correction for multiple comparisons with a threshold of p <0.05, with age and sex as covariates.

## Results

### Participants

108 participants with PD were included (all from the UCL site); 61 people with DLB were included, with 36 from the UCL site, and 25 from the NACC site. A total of 165 controls were included, 38 from UCL and 127 from the NACC database (ADRC “8361”), these control participants were used to calibrate the datasets to the Rutherford and colleagues’ reference dataset (n=58,836) [12]. Age and sex for respective groups and differences between them are shown in **Table 1**. We adjusted for age and sex in subsequent analyses and also note that an important strength of the neuroanatomical normative modelling pipeline is that it has been modelled to account for age, sex and site differences.

**Table 1.** Demographics and total outlier counts.

Overall PD vs DLB
|  | PD (n=108) | DLB (n=61) | Statistic |
| --- | --- | --- | --- |
| Mean age (SD), y | 64.1 (7.8) | 73.8 (6.5) | **t=-9.2; P<0.01** |
| Male, n (%) | 51 (48) | 55 (90) | **$\chi^2=27.9$; P<0.01** |
| Mean tOC (SD) | 3.6 (6.0) | 8.7 (11.3) | **$\beta= -5.60$ (SE=1.74); P<0.01\*** |

High vs Low visual Performers with PD
|  | High (n=62) | Low (n=34) | Statistic |
| --- | --- | --- | --- |
| Mean age (SD), y | 61.5 (7.1) | 68.1 (8.0) | **t=-4.02; P<0.01** |
| Male, n (%) | 31 (50) | 14 (41) | $\chi^2=0.38$; P=0.54 |
| Mean tOC (SD) | 2.3 (3.5) | 4.1 (5.2) | **$\beta= -4.73$ (SE=1.30); P<0.01\*** |

DLB by site (UCL vs NACC participants)
|  | UCL (n=36) | NACC (n=25) | Statistic |
| --- | --- | --- | --- |
| Mean age (SD), y | 72.9 (5.5) | 75.1 (6.3) | t=-1.41; P=0.17 |
| Male, n (%) | 33 (92) | 22 (88) | $\chi^2=0.001$; $P=0.97$ |
| Mean tOC (SD) | 6.3 (9.3) | 12.1 (13.2) | **$\beta= -6.02$ (SE=3.00); $P=0.048^*$** |
| PD vs DLB at UCL site only |  |  |  |
|  | PD (n=108) | DLB (n=36) | Statistic |
| Mean age (SD), y | 64.1 (7.8) | 72.9 (5.5) | **$t= -7.54$; $P<0.01$** |
| Male, n (%) | 51 (48) | 33 (92) | **$\chi^2=19.4$; $P<0.01$** |
| Mean tOC (SD) | 3.6 (6.0) | 6.3 (9.3) | $\beta= -3.13$ (SE=1.61); $P=0.054^*$ |
| PD, Parkinson's disease; DLB, Dementia with Lewy bodies; tOC, total outlier count; UCL, University College London; NACC, National Alzheimer's Co-ordinating Centre |  |  |  |
| *P values were analysed by a linear regression adjusting for age and sex |  |  |  |
| **BOLD** signifies statistically significant difference |  |  |  |

### Differences in total outlier count between DLB and PD

Mean total outlier count was significantly higher in the overall DLB group (n=61; mean=8.7 (SD=11.3)) compared to the PD group (n=108; mean=3.60 (SD=6.0)) after adjusting for age and sex (PD vs DLB: β= −5.60 (SE=1.74); t=-3.23; P<0.01). It was also higher in the low (n=34; mean=5.4 (SD=8.7)) compared to high (n=62; mean=2.3 (SD=3.5)) visual performer group after adjusting for age and sex (β= −4.73 (SE=1.30); t=-3.64; P<0.01). The DLB participants from the NACC site (n=25) had significantly higher numbers of outliers compared to those from the UCL site (n=36) after adjustment for age and sex (UCL vs. NACC: β= −6.02 (SE=2.97); t=-2.02; P=0.050). At the UCL site alone, there was a numerically higher number of outliers in the DLB (n=36) compared to PD group (n=108) but this did not reach statistical significance (PD vs DLB at UCL site: β= −3.13 (SE=1.61); t= −1.95; P=0.054) (**Table 1**).

### Heterogeneity in patterns of outliers found between PD at-risk of dementia groups and between DLB and PD

Dissimilarity as measured by individual median Hamming distance scores metric was significantly higher in PD participants who were low visual performers (n=34; mean=7.1 (SD=8.4)) compared to high visual performers (n=62; mean=3.3 (SD=3.4), W=522.5; P<0.01). The Hamming distance matrices for each group and a density plot of Hamming distances are shown in **Figure 1A-C**.

**Figure 1.**
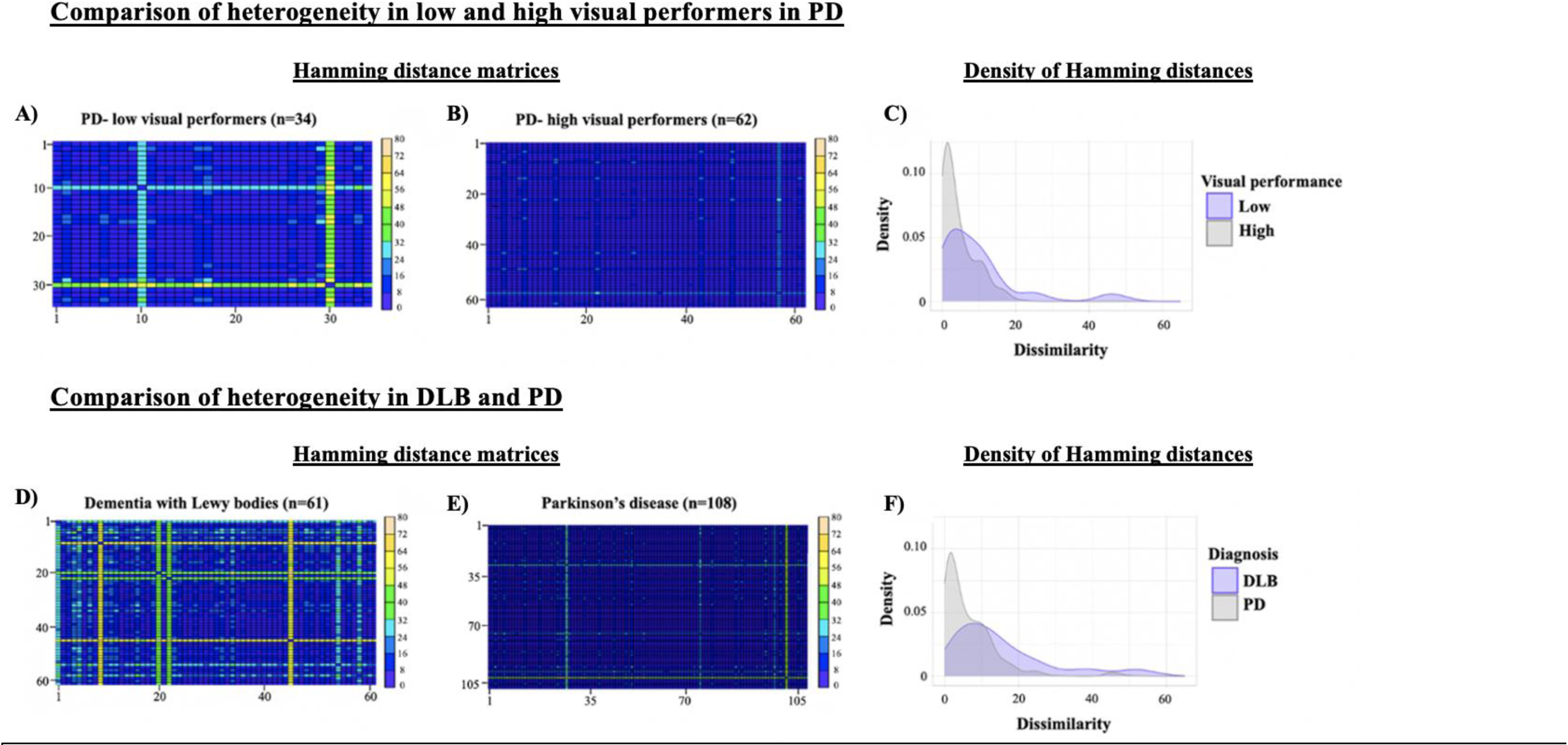
Outlier Heterogeneity. Outlier Hamming distance matrices for PD-low visual performers (A) and PD-high visual performers (B). Kernel density estimates (Y-axis) for a given Hamming distance score (X-axis) show that PD-low visual performers had more dissimilarity as evidenced by the shorter peak and longer tail compared to PD-high visual performers (C). Outlier Hamming distance matrices for the DLB (D) and PD (E) groups. Kernel density estimates (Y-axis) for a given Hamming distance score (X-axis) show that DLB participants had more dissimilarity as evidenced by the shorter peak and longer tail compared to the overall PD group (F). In A, B, D, E: dark blue / indigo represents the lower end of hamming distance scores whereby two participants are relatively similar to one another in terms of regional distribution of outliers whereas yellow represents higher hamming distance scores signifying greater dissimilarity. The more yellow in the plot, the greater the dissimilarity between individuals in the groups. PD, Parkinson’s disease; DLB, Dementia with Lewy bodies

The individual median Hamming distances score was significantly higher in DLB (n=61; mean=12.6 (SD=10.3)) compared to PD (n=108; mean=4.6 (SD=6.0), (W=5649; P<0.01). The Hamming distance matrices for DLB and PD, and a density plot of Hamming distances is shown in **Figure 1D-F**.

The proportion of regional outliers were mapped by group (**Figure 2**) and indicated that for low compared to high visual performers with PD, and for DLB compared to PD, there are increased number of regions in which there are outliers, suggesting greater heterogeneity and more widespread atrophy.

**Figure 2.**
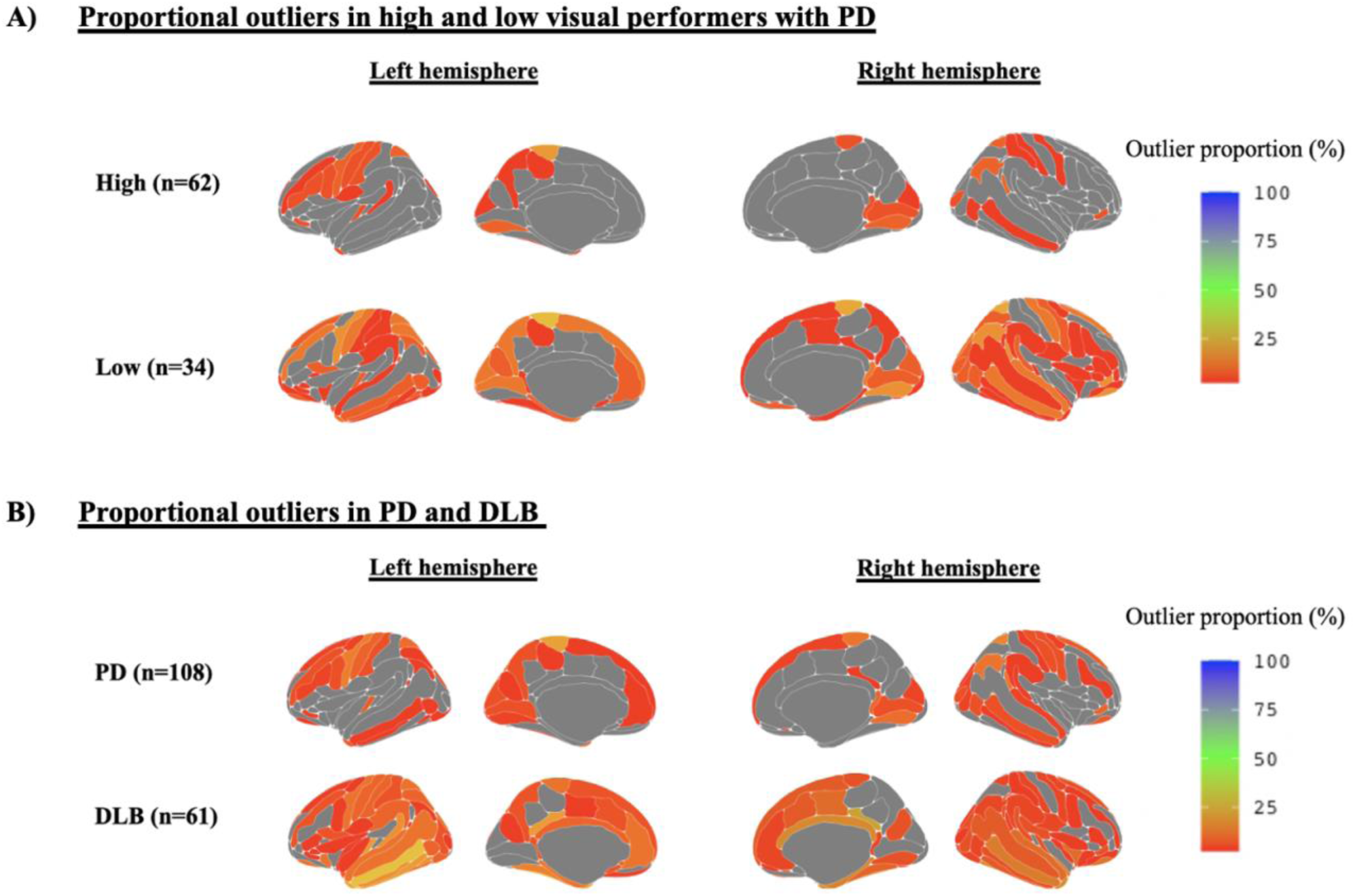
Regional maps of outliers. Mapped are the proportion of participants who are outliers in a particular cortical region. A. Low visual performers (who are at-risk of Parkinson’s dementia) compared to high visual performers with PD (at lower risk of Parkinson’s dementia). Qualitatively, more regions have a higher proportion of participants with outliers in the low visual performer group. B. PD and DLB. Qualitatively, more regions have higher proportions of participants with outliers in DLB than PD. Of note, there is no one region with more than 25% of participants being outliers, highlighting the heterogeneity in cortical atrophy in DLB and PD.. Grey represents regions with 0-2.5% outliers.

In the low visual performers PD group, 99 out of the 169 brain regions showed at least one patient with an outlier. This compares with 77/169 regions having at least one outlier in the high visual performers group. The maximum number of participants who were an outlier in any one region was 6 (12.9%) for the high visual performers and 8 (17.6%) for low visual performers, both being in the in left *Paracentral lobule and sulcus* region (**Supplementary Table 1**). Further information on the proportion of outliers in each region, and where significant regional differences between groups exist, are shown in the supplementary material (**Supplementary Table 1; Supplementary Figure 1**).

In the PD group as a whole, 125 regions out of 169 had at least one patient with an outlier. This compares with 147/169 regions in the DLB group. The region with the highest number of participants who were an outlier was the left Paracentral lobule and sulcus region (n=15, 13.9%). For the DLB group the region with the highest number of outliers was the right Posterior-dorsal part of the cingulate gyrus (dPCC), which had 15 people (24% of the group) with outlier scores for this region.(**Supplementary Table 1; Supplementary Figure 1**).

### Total outlier counts are associated with cognitive performance in DLB and with visuospatial processing in PD

There were significant differences in several clinical measures, including composite cognitive score, MoCA, Hooper Visual Organisation Test, MDS-UPDRS, MDS-UPDRS motor symptom subscale, UM-PDHQ and HADS depression scores, between PD and DLB groups, with the latter more severely affected **(Table 2).**

**Table 2.** Clinical Features of PD and DLB groups from UCL site.

| **Table 2. Clinical Features of PD and DLB groups from UCL site** |  |  |  |
| --- | --- | --- | --- |
|  | **PD (n=108)** | **DLB (n=36)** | **Statistic** |
| **Age, y** | 64.1 (7.8) | 72.5 (5.6) | **t=6.76; P<0.01** |
| **Male, n (%)** | 51 (48) | 29 (91) | **$\chi^2=16.89$; P<0.01** |
| **Disease Duration (years)** | 4.1 (2.5) | 3.7 (2) | W=2103, P=0.46 |
| **Composite cognitive score** | -0.27 (0.76) | -2.93 (1.88) | **W=268; P<0.01** |
| **MoCA** | 28.0 (1.9) | 20.8 (5.8) | **W=385.5; P<0.01** |
| **HVOT** | 24.4 (3.1) | 16.4 (6.5) | **W=476; P<0.01** |
| **MDS-UPDRS** | 45.5 (21.4) | 66.6 (27.5) | **W=2953; P<0.01** |
| **MDS-UPDRS motor** | 19.7 (13.6) | 34.9 (15.7) | **W=3075.5; P<0.01** |
| **UM-PDHQ** | 0.7 (1.8) | 4.3 (3.0) | **W=3250; P<0.01** |
| **HADS depression** | 4.0 (2.9) | 5.7 (3.2) | **W=2553; P<0.01** |
| All data shown are mean (SD) except sex. |  |  |  |
| Significant differences are highlighted in **bold**. |  |  |  |
| PD, Parkinson's disease; DLB, Dementia with Lewy bodies; MoCA, Montreal Cognitive Assessment; Hooper Visual Organisation Test, Hooper Visual Organisation Test; HADS, Hospital Anxiety and Depression Scale; MDS-UPDRS, Movement Disorders Society Unified Parkinson's Disease Rating Scale; UM-PDHQ, University of Miami Hallucinations; Questionnaire; HADS, Hospital Anxiety and Depression Scale |  |  |  |

In DLB, there were significant associations between greater total outlier count and worse performance in cognitive scores, with both the composite cognitive score (β=-2.01 (SE=0.79); t=-2.54; P=0.02) and MoCA (β=-0.55 (SE=0.27), t=-2.04, P=0.05), when covarying for age and sex. However, there was no significant association with a test of visuoperceptual ability, the Hooper Visual Organisation Test (β=-0.45 (SE=0.24); t=-1.89; P=0.068) (**Figure 3)**. There were also no significant associations with other measures of Lewy body disease severity (MDS-UPDRS score), motor symptoms (MDS-UPDRS-III score), hallucination severity (UM-PDHQ) or depression (HADS depression score) (**Table 3**).

**Figure 3.**
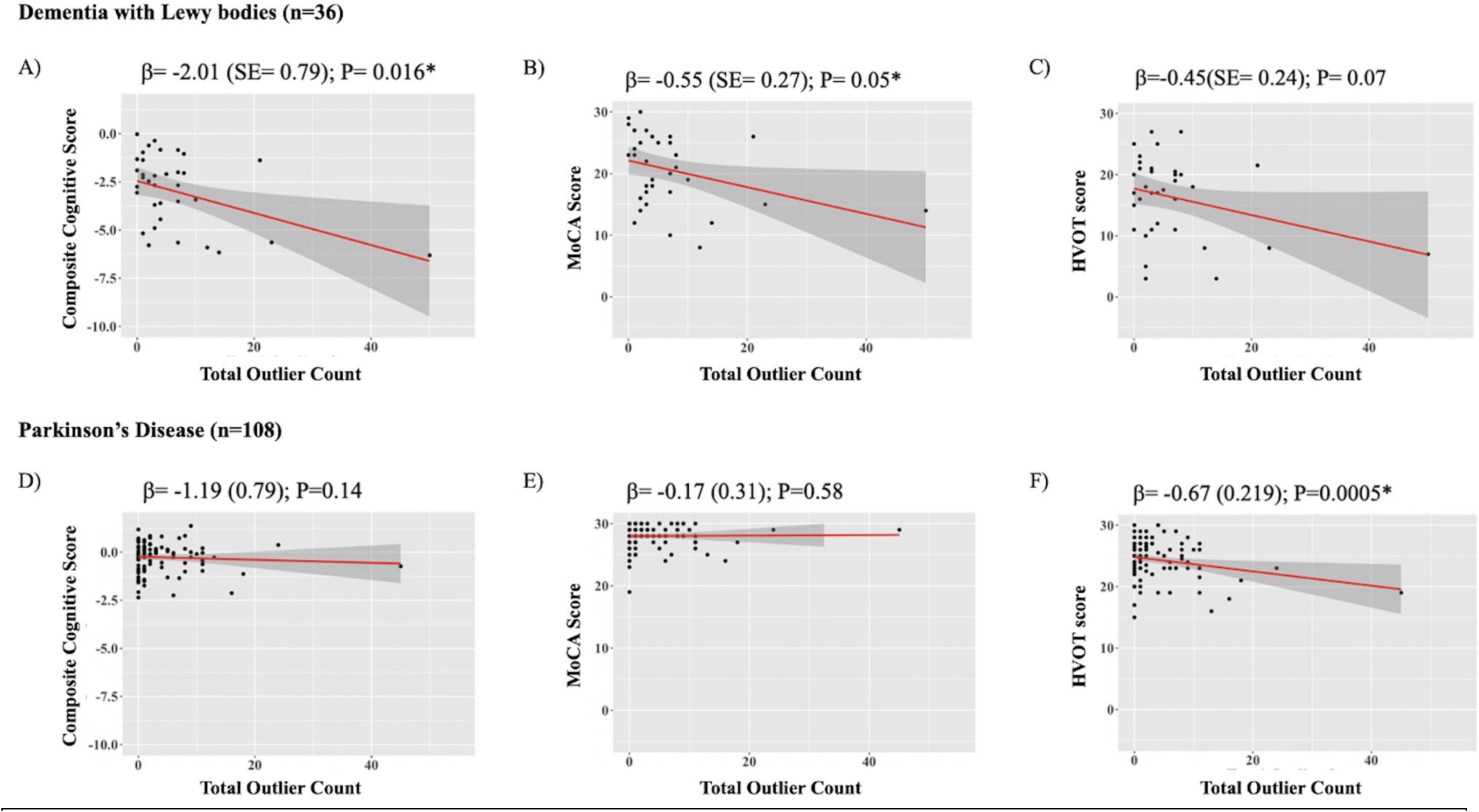
Relationship between total outlier count and cognitive measures in DLB and PD. Regression plots for the association between total outlier count (independent variable) and the following dependent variables: Composite Cognitive score, MoCA and Hooper Visual Organisation Test, in DLB (A, B, C, respectively) and PD (D, E, F, respectively). β coefficient values, corrected for age and sex, are presented along with P values. * denotes significant association. MoCA, Montreal Cognitive Assessment; HVOT, Hooper Visual Organisation Test; DLB, Dementia with Lewy bodies; PD, Parkinson’s disease.

**Table 3.** Association of total outlier count with measures of cognitive performance and other disease specific measures.

| Attribute | PD (n=108) |  |  |  | DLB (n=36) |  |  |  |
| --- | --- | --- | --- | --- | --- | --- | --- | --- |
|  | beta | SE | t | P value<sup>a</sup> | beta | SE | t | P value<sup>a</sup> |
| Cognitive performance |  |  |  |  |  |  |  |  |
| Composite Cognitive Score | -1.19 | 0.79 | -1.51 | **0.14** | **-2.01** | **0.79** | **-2.54** | **0.016** |
| MoCA | -0.17 | 0.31 | -0.55 | **0.58** | **-0.55** | **0.27** | **-2.04** | **0.050** |
| HVOT | **-0.67** | **0.19** | **-3.59** | **<0.01** | -0.45 | 0.24 | -1.89 | 0.068 |
| Disease-specific measures |  |  |  |  |  |  |  |  |
| MDS-UPDRS | 0.03 | 0.03 | 1.01 | 0.31 | 0.03 | 0.03 | 0.74 | 0.47 |
| MDS-UPDRS Motor Score | -0.05 | 0.04 | -1.18 | 0.24 | 0.06 | 0.06 | 0.95 | 0.35 |
| UM-PDHQ | 0.44 | 0.32 | -1.38 | 0.17 | 0.30 | 0.53 | 0.56 | 0.58 |
| HADS depression | 0.05 | 0.19 | 0.26 | 0.80 | -0.39 | 0.32 | -1.23 | 0.23 |

PD, Parkinson's disease; DLB, Dementia with Lewy bodies; MoCA, Montreal Cognitive Assessment; HVOT, Hooper Visual Organisation Test; HADS, Hospital Anxiety and Depression Scale; MDS-UPDRS, Movement Disorders Society Unified Parkinson's Disease Rating Scale; UM-PDHQ, University of Miami Hallucinations; Questionnaire; HADS, Hospital Anxiety and Depression Scale.
<sup>a</sup>P values were analysed using linear regressions adjusting for age and sex.
In **bold** results showing statistically significant associations

In the PD group, the only significant association was with visuoperceptual ability, the Hooper Visual Organisation Test (β=-0.67 (SE=0.19); t=-3.59; P<0.01), with no association seen for the other cognitive scores. Similar to DLB, no associations were found in PD between other disease measures and total outlier count.

In the PD and DLB groups, one participant in each group had a markedly larger number of total outliers than the other participants in the same group, with 45 and 50 total outliers respectively. Though biologically plausible and not erroneously measured, we performed an additional sensitivity analysis by testing the association of total outlier counts with cognitive measures without these individuals included. In the DLB group, the effects of greater total outlier count with poorer composite cognitive score ((β=-0.94 (SE=0.51); t=-1.83; P=0.078) and MoCA (β=-0.33 (SE=0.16); t=-2.01; P=0.053) were weakened, and were now below the level of statistical significance. In the PD group, the significant association with the Hooper Visual Organisation Test (β=-0.38 (SE=0.15); t=-2.59; P=0.011) remained (**Supplementary Figure 2**).

### Group-level cortical thickness analysis is less sensitive to differences in cortical atrophy between groups

A conventional GLM approach using Monte Carlo correction for multiple comparisons, thresholded to significance level of P<0.05, did not find any significant clusters of differences in cortical thickness between high and low visual performers in PD. Comparing the overall PD group with DLB, there were two significant clusters in the left precentral region and one significant cluster in both the superior frontal and precentral regions on the right, signifying reduced cortical thickness in DLB compared with PD in these regions (**Supplementary Table 2)**.

## Discussion

We used neuroanatomical normative modelling to examine heterogeneity in brain atrophy at the level of the individual in PD and DLB to reveal new insights into patterns of brain atrophy in these conditions. Strikingly, we found limited overlap of outlier patterns in both PD and DLB, with increased inter-individual dissimilarity and more widespread outlier patterns in low compared to high visual performers with PD, and in DLB compared to the overall PD group. We also found that the total outlier count was significantly higher in patients at higher risk of developing dementia (revealed by low visual performance), compared to PD patients who were high visual performers (and therefore at low risk of dementia). Patients with DLB showed higher total outlier counts than patients with PD overall. Finally, total outlier count was significantly associated with severity of clinically relevant cognitive measures in both PD and DLB. Overall, this suggests that the total outlier count derived from neuroanatomical normative modelling may have utility as a novel neuroimaging measure in Parkinson’s and DLB.

Importantly, we were able to detect differences in the total outlier count, especially between dementia risk groups in Parkinson’s, where more conventional group-level GLM analysis of regional cortical thicknesses could not detect group differences. This is because the total outlier count is agnostic to the regional location of cortical atrophy, whereas GLM approaches require cortical atrophy to be in similar locations between individuals. In conditions such as Parkinson’s and Lewy Body Disorders, where there are greater inter-individual differences in patterns of atrophy [3, 41], the total outlier count is likely to be a more sensitive measure of overall cortical atrophy.

We were able to detect these differences in total outlier count in patients at different stages in progression to dementia, in a Parkinson’s at-risk group (where patients did not yet have dementia); as well as in DLB. This suggests that measures such as the total outlier count, and approaches such as neuroanatomical normative modelling may have clinical utility as a prognostic neuroimaging measure of the risk of disease progression in Lewy body disorders, as has been shown in Alzheimer’s disease previously [13, 14]. Importantly for its potential application in clinical contexts, the total outlier count can be calculated based on cortical thicknesses and subcortical volume read-outs generated by applying an automated software pipeline, available in the FreeSurfer package, to structural T1w-MRI data. Total outlier count has the added potential of being a translational, cost-effective, and generalisable neuroimaging measure as T1w-MRI is routinely collected in clinical practice.

Higher total outlier count was significantly associated with lower composite cognitive score and the MoCA in DLB but not a lower score on a measure of visuoperceptual processing. In contrast, in PD, we did not find a relationship between cognitive scores and the total outlier count; whereas we did find a relationship between total outlier count and visuoperception. It is possible that the lack of a linear relationship between cognitive measures and the total outlier count in PD was due to ceiling effects in the MoCA and combined cognitive scores. On the other hand, the Hooper test of visual organisation, which measures visuoperception, may be a particularly sensitive measure of cognitive impairment in PD because visuoperceptual ability (together with visuospatial deficits) are early and key cognitive domains affected in PD [42, 43]. As a result, this measure may be less prone to ceiling effects. Our findings are in keeping with previous work that showed an association of higher total outlier count with worse cognitive performance in memory and executive functioning domains across participants with AD and mild cognitive impairment as well as healthy controls [14].

There was increased inter individual heterogeneity in low compared to high visual performers with PD, and in DLB compared to PD. In both comparisons, the group associated with poorer cognitive functioning showed increased heterogeneity. This is consistent with previous work that found increased heterogeneity in Alzheimer’s compared to mild cognitive impairment and controls [14]. The differences between PD and DLB may relate to greater cortical involvement in DLB [44]. Our work is the first to objectively quantify inter individual heterogeneity in atrophy measures in PD and DLB and highlights the challenges associated with interpretation of group level analyses in neurodegenerative disorders which fail to address this.

There are some limitations associated with this work. Firstly, the reference normative model does not factor in biological differences in neuroanatomy influenced by ethnicity, and we have not adjusted for this in our analyses as this data was not available. This limits the generalisability of our findings to broader populations. An aspiration for the development of future reference normative models will be to better capture intrinsic biological differences in brain atrophy driven by ethnicity. Secondly, whilst the total outlier count metric has potential utility as a neuroimaging biomarker, outliers were calculated as cortical thickness z-scores <-1.96. Therefore, this metric may fail to capture lesser degrees of cortical neurodegeneration which are potentially relevant but do not reach the threshold required to be classed as an outlier. A further limitation is that there are inherent differences between sites from which DLB participant data was collected. The data from the UCL site was collected in a bespoke prospective fashion with our study aims in mind, whereas the NACC data is from a large relational database. This meant that some relevant data for our work relating to neuropsychological testing and clinical features were unavailable. Though there were no significant age and sex differences between these subgroups, the participants from the NACC site had significantly increased total outlier counts, which may suggest a more advanced disease stage. This may in part be due to differences in study inclusion criteria between sites, as well as testing demands on participants at the UCL site, leading to possible selection bias of those who are less functionally impaired at the UCL site.

Future work should also be adequately powered to capture individual level differences in DLB subgroups and to investigate whether there are different patterns of heterogeneity and total outlier counts between such subgroups. This could include, for example, DLB patients who are depressed versus non-depressed. Additionally, longitudinal data would enable individual z-score trajectories to be evaluated over time and this could be crucial in confirming the utility of the total outlier count as a prognostic measure; and as a measure of disease progression.

## Conclusion

We used neuroanatomical normative modelling to examine differences in brain atrophy patterns between PD patients with good and poor vision, who are in differing risk groups for PD dementia, and between patients with PD and DLB. We showed that PD patients with poor visual function have a greater degree of loss of cortical thickness and subcortical volumes, measured as total outlier counts; and that patients with DLB had greater total outlier count than those with PD. We further showed that total outlier count was significantly associated with global cognitive performance in DLB; and with visuoperception in PD. Neuroanatomical normative modelling is therefore a useful approach for degenerative conditions such as PD and DLB with more variable patterns of atrophy and one of the key metrics that is derived, the total outlier count, holds significant promise as a clinically useful measure of disease progression.

## Author Contributions

Rohan Bhome, Rimona S Weil and James H Cole conceived the study. Rohan Bhome, Ivelina Dobreva and Naomi Hannaway collected data. Rohan Bhome, Serena Verdi, Sophie A Martin, Neil P Oxtoby and Gonzalo Castro Leal contributed to data processing and statistical analysis. Rohan Bhome wrote the first draft of the manuscript and all authors edited and agreed to the final version of the manuscript.

## Data Availability

All data produced in the present study are available upon reasonable request to the authors

## Acknowledgements

The authors thank all the participants for their time.

Rohan Bhome. is supported by a Wolfson-Eisai Clinical Research Training Fellowship. Naomi Hannaway is supported by a grant by the Rosetrees and Stoneygate Trusts. Neil P Oxtoby and Gonzalo Castro Leal acknowledge support from a UKRI Future Leaders Fellowship (MR/S03546X/1) and the National Institute for Health Research University College London Hospitals Biomedical Research Centre. A.F. Marquand gratefully acknowledges funding from the Dutch Organization for Scientific Research via a VIDI fellowship (grant number 016.156.415). Rimona S Weil is supported by a Wellcome Clinical Research Career Development Fellowship (205167/Z/16/Z).

The NACC database is funded by NIA/NIH Grant U24 AG072122. NACC data are contributed by the NIA-funded ADRCs: P30 AG062429 (PI James Brewer, MD, PhD), P30 AG066468 (PI Oscar Lopez, MD), P30 AG062421 (PI Bradley Hyman, MD, PhD), P30 AG066509 (PI Thomas Grabowski, MD), P30 AG066514 (PI Mary Sano, PhD), P30 AG066530 (PI Helena Chui, MD), P30 AG066507 (PI Marilyn Albert, PhD), P30 AG066444 (PI John Morris, MD), P30 AG066518 (PI Jeffrey Kaye, MD), P30 AG066512 (PI Thomas Wisniewski, MD), P30 AG066462 (PI Scott Small, MD), P30 AG072979 (PI David Wolk, MD), P30 AG072972 (PI Charles DeCarli, MD), P30 AG072976 (PI Andrew Saykin, PsyD), P30 AG072975 (PI David Bennett, MD), P30 AG072978 (PI Neil Kowall, MD), P30 AG072977 (PI Robert Vassar, PhD), P30 AG066519 (PI Frank LaFerla, PhD), P30 AG062677 (PI Ronald Petersen, MD, PhD), P30 AG079280 (PI Eric Reiman, MD), P30 AG062422 (PI Gil Rabinovici, MD), P30 AG066511 (PI Allan Levey, MD, PhD), P30 AG072946 (PI Linda Van Eldik, PhD), P30 AG062715 (PI Sanjay Asthana, MD, FRCP), P30 AG072973 (PI Russell Swerdlow, MD), P30 AG066506 (PI Todd Golde, MD, PhD), P30 AG066508 (PI Stephen Strittmatter, MD, PhD), P30 AG066515 (PI Victor Henderson, MD, MS), P30 AG072947 (PI Suzanne Craft, PhD), P30 AG072931 (PI Henry Paulson, MD, PhD), P30 AG066546 (PI Sudha Seshadri, MD), P20 AG068024 (PI Erik Roberson, MD, PhD), P20 AG068053 (PI Justin Miller, PhD), P20 AG068077 (PI Gary Rosenberg, MD), P20 AG068082 (PI Angela Jefferson, PhD), P30 AG072958 (PI Heather Whitson, MD), P30 AG072959 (PI James Leverenz, MD).

## Abbreviated title

Normative modelling in Parkinson’s disease and Dementia with Lewy bodies

