## Supplementary for "A neuroimaging measure to capture heterogeneous patterns of atrophy in Parkinson’s disease and dementia with Lewy bodies"

**Supplementary Material**

**Title**

### Neuroanatomical normative modelling in Parkinson's disease and Dementia with Lewy bodies unveils a novel neuroimaging biomarker

**Authors**

Bhome R[1,2], Verdi S[1,2], Martin SA[2], Hannaway N[1], Dobreva I[1], Oxtoby NP[2], Casto-Leal G, Rutherford S[5,6,7] Marquand AF[5,6,8], *Weil RS[1,3,4], *Cole JH[1,2]

| Supplementary Table 1. Brain regions with highest percentage of outliers in each study group | | | |
|---|---|---|---|
| **Brain region** | **Number (%) of outliers** | **Brain region** | **Number (%) of outliers** |
| **PD- high visual performers (n=62)** | | **PD- low visual performers (n=34)** | |
| Left Paracentral lobule and sulcus | 8 (12.9) | Left Paracentral lobule and sulcus | 6 (17.7) |
| Right Superior parietal lobule | 4 (6.5) | Right Superior parietal lobule | 5 (14.7) |
| Right Lingual gyrus | 4 (6.5) | Right Paracentral lobule and sulcus | 5 (14.7) |
| Right Angular gyrus | 4 (6.5) | Right Lingual gyrus | 5 (14.7) |
| Left Precentral gyrus | 4 (6.5) | Right Angular gyrus | 5 (14.7) |
| Left Superior parietal lobule | 4 (6.5) | Left Precentral gyrus | 5 (14.7) |
| Left Transverse frontopolar gyri and sulci | 4 (6.5) | Right Orbital part of the inferior frontal gyrus | 5 (14.7) |
| Left Lingual gyrus | 4 (6.5) | Right Orbital sulci | 5 (14.7) |
| **PD (n=108)** | | **DLB (n=61)** | |
| Left Paracentral lobule and sulcus | 15 (13.9) | Right dPCC | 15 (24.6) |
| Right Superior parietal lobule | 11 (10.9) | Left Lateral occipito-temporal gyrus | 12 (19.7) |
| Left Precentral gyrus | 10 (9.3) | Left Middle temporal gyrus | 11 (18.0) |
| Right Paracentral lobule and sulcus | 9 (8.3) | Right Temporal pole | 11 (18.0) |
| Right Angular gyrus | 9 (8.3) | Right Lateral occipito-temporal sulcus | 11 (18.0) |
| | | Right Pericallosal sulcus | 11 (18.0) |

Top 5 regions in terms of percentage of outliers for each study group. Where there is a tie, all regions are shown.

PD, Parkinson’s disease; DLB, Dementia with Lewy bodies; dPCC, Posterior-dorsal part of the cingulate gyrus

**Supplementary Table 2. Significant clusters for the left and right hemisphere comparing PD and DLB cortical thicknesses.**

| | Region | Cluster size ($mm^2$) | MNI Coordinates | | | CWP |
|---|---|---|---|---|---|---|
| | | | x | y | z | |
| **Left hemisphere** | | | | | | |
| | Precentral | 103.33 | -29.5 | -13.1 | 58.9 | 0.013 |
| | Precentral | 90.01 | -33.7 | -16.1 | 41.1 | 0.023 |
| **Right hemisphere** | | | | | | |
| | Superiorfrontal | 110.67 | 16.4 | -3.5 | 66.7 | 0.011 |
| | Precentral | 89.84 | 30.7 | -12.4 | 52.0 | 0.020 |

P values from the Monte carlo simulation and clustering as cluster wise probability (CWP), resulting from the vertex-wise comparison of cortical thickness between PD and DLB participants.

MNI, Montreal Neurological Institute; PD, Parkinson’s disease; DLB, Dementia with Lewy bodies

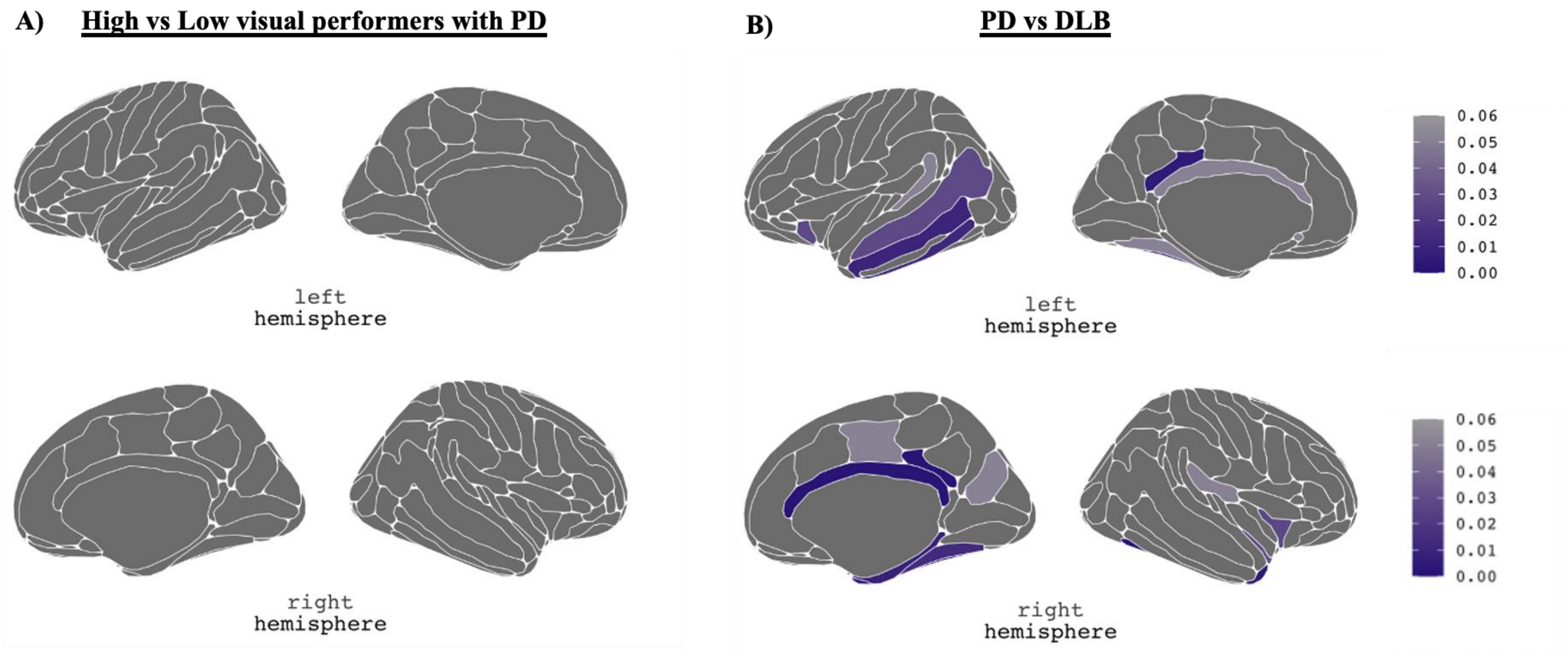


**Supplementary Figure 1. Regional maps of group differences in proportional outliers.** There were no FDR corrected significant differences in the proportion of outliers in any cortical region between high and low visual performers with PD (A); however there were sixteen regions where there were FDR corrected significant differences (FDR P value <0.05) when comparing PD with DLB, and in each case the proportion of outliers was greater for DLB: right dPCC, right lateral occipito-temporal sulcus, right pericallosal sulcus, left dPCC, right temporal pole, left lateral occipito-temporal gyrus, right parahippocampal gyrus, right lateral occipito-temporal gyrus, left middle temporal gyrus, left inferior temporal gyrus, right medial occipito-temporal sulcus and lingual sulcus, right short insular gyri, right hippocampus, left superior temporal sulcus,left anterior segment of the circular sulcus of the insula, right planum polare of the superior temporal gyrus

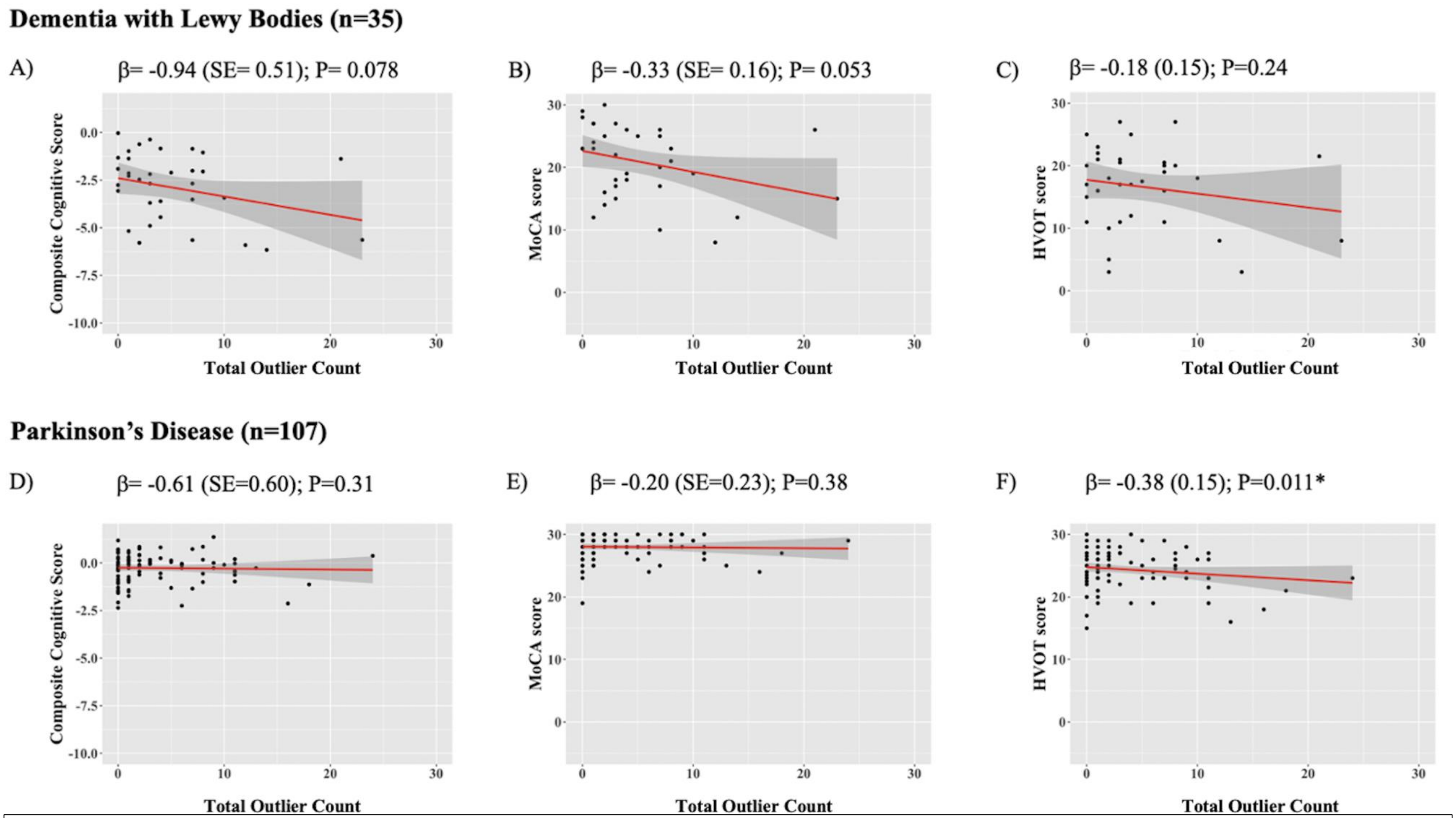


**Supplementary Figure 2. Relationship between total outlier count, with potential outlying scores excluded, and cognitive measures in DLB and PD.** Regression plots for the association between total outlier count (independent variable) and the following dependent variables: Composite Cognitive score, MoCA and HVOT, in DLB (A, B, C , respectively) and PD (D, E, F, respectively).

β coefficient values, corrected for age and sex, are presented along with P values. * denotes significant association.

MoCA, Montreal Cognitive Assessment; HVOT, Hooper Visual Organisation Test; DLB, Dementia with Lewy bodies; ; PD,. Parkinson's disease
